# High mortality of COVID-19 associated mucormycosis in France: a nationwide retrospective study

**DOI:** 10.1101/2021.07.05.21260041

**Authors:** François Danion, Valérie Letscher-Bru, Juliette Guitard, Karine Sitbon, Sarah Dellière, Adela Angoulvant, Guillaume Desoubeaux, Francoise Botterel, Anne-Pauline Bellanger, Gilles Gargala, Fabrice Uhel, Marie-Elisabeth Bougnoux, Victor Gerber, Justin Michel, Marjorie Cornu, Stéphane Bretagne, Fanny Lanternier, the COVID-Mucor study group

## Abstract

We studied COVID-19 associated mucormycosis based on 17 cases reported nationwide and assessed the differences with India. They differed by frequencies of diabetes mellitus (47% in France versus 95% in India), hematological malignancies (35% versus 1%), anatomical sites (53% lung versus >80% rhino-orbito-cerebral) and prognosis (>80% mortality versus <50%).

## INTRODUCTION

Coronavirus disease 2019 (COVID-19) has a wide spectrum of severity. Fungal superinfections, notably aspergillosis, can complicate the course of severe COVID-19 with a high mortality [1]. Emerging reports, mainly from India, recently described COVID-19-associated mucormycosis (CA-Mucor). In this country, more than 28,000 cases have already been reported and mucormycosis is now a notifiable disease [2]. Outside India, only few case reports have been published from some countries including Europe [3,4]. The aim of our study was to describe cases of COVID-19 associated mucormycosis (CA-Mucor) in France and analyze host factors, presentation and outcome.

## METHODS

We conducted a retrospective nationwide study on CA-Mucor. Our network of 59 French mycology laboratories were requested to report CA-Mucor cases diagnosed from March 2020 to June 2021 to the French National Reference Center for Invasive Mycosis & Antifungals (NRCMA) as part of its surveillance missions. Only cases occurring within the 3 months after COVID-19 diagnosis confirmed by a positive PCR for SARS-CoV-2 were included. Clinical data were recorded anonymously on a standardized case report form. Cases were classified as proven or probable according to EORTC/MSGERC criteria [5], with the addition of diabetes mellitus and dexamethasone prescribed for COVID-19 as host factor and positive *Mucorales* PCR in serum, blood or plasma as mycological evidence. Date of mucormycosis diagnosis was defined by the first positive sample for mucormycosis. This study is part of the NRCMA official duties which are approved by the Institut Pasteur Internal Review Board (2009-34/IRB) in accordance with French Law. One case has already been published [6].

## RESULTS

Seventeen patients from eleven centers developed CA-Mucor. Sixteen (94%) patients were male and the median age was 64 [range 25 – 79]. The median body mass index was 28 [range 19 – 37].

### Underlying risk factors before COVID-19

Among the seventeen patients, seven (41%) had classical EORTC/MSGERC host factors for invasive mold infections before COVID-19, including one with solid organ transplantation and six (35%) with hematological malignancies (HM) [5]. Among the latter, four (24%) had neutropenia within the last month, one had allogeneic and one autologous hematopoietic stem cell transplant. Overall, four (24%) patients had diabetes mellitus (DM), including one with HM. Seven (41%) patients had no risk factors for mucormycosis before COVID-19, but one had pulmonary carcinoma and one underwent chronic dialysis. Three (18%) patients received immunosuppressive drugs, two (12%) long-term corticosteroids, and two (12%) were currently having antineoplastic chemotherapy.

### COVID-19

Sixteen (94%) patients had severe COVID-19 requiring intensive care unit (ICU). Median time between first COVID-19 symptoms and ICU transfer was 7 days [range 0 – 86]. Management of COVID-19 required corticosteroids for thirteen (76%) patients, mainly dexamethasone, and tocilizumab for two (12%). To note, one patient received ≥0.3 mg/kg corticosteroids for ≥3 weeks. Twelve (71%) patients had high flow nasal cannula oxygen therapy, and thirteen (76%) had invasive mechanical ventilation. Only two patients had neither high flow nasal cannula oxygen therapy nor mechanical ventilation.

Four (24%) patients developed DM induced by dexamethasone for COVID-19, meaning that overall eight patients (47%) had DM. Four (24%) patients had diabetic ketoacidosis. Eleven (65%) patients had renal failure, eight (47%) requiring dialysis.

### Mucormycosis

CA-Mucor was diagnosed a median of 24 days [range 8 – 90] after COVID-19 first symptoms, 12.5 days [range 1 – 49] after ICU hospitalization and 16 days [range 1 – 49] after corticosteroid prescription.

CA-Mucor location was mainly pulmonary (n=9; 53%), but also digestive (n=3; 18%), rhino-orbito-cerebral (n=2; 12%) or disseminated (n=3; 18%). The six patients with HM had pulmonary (n=3) or disseminated (n=3) CA-Mucor. The rhino-orbito-cerebral forms occurred in one patient with DM and one patient with lung transplant. The three digestive CA-Mucor occurred in patients with no host factors, including no dexamethasone-induced DM.

Twelve patients with a pulmonary location had a chest CT scan evidencing a reversed halo sign in one patient (8%) with HM and neutropenia (Figure 1A), consolidation in ten (83%) patients, including four (33%) with a cavitation (Figure 1B), and one with nodules.

**Figure 1.**
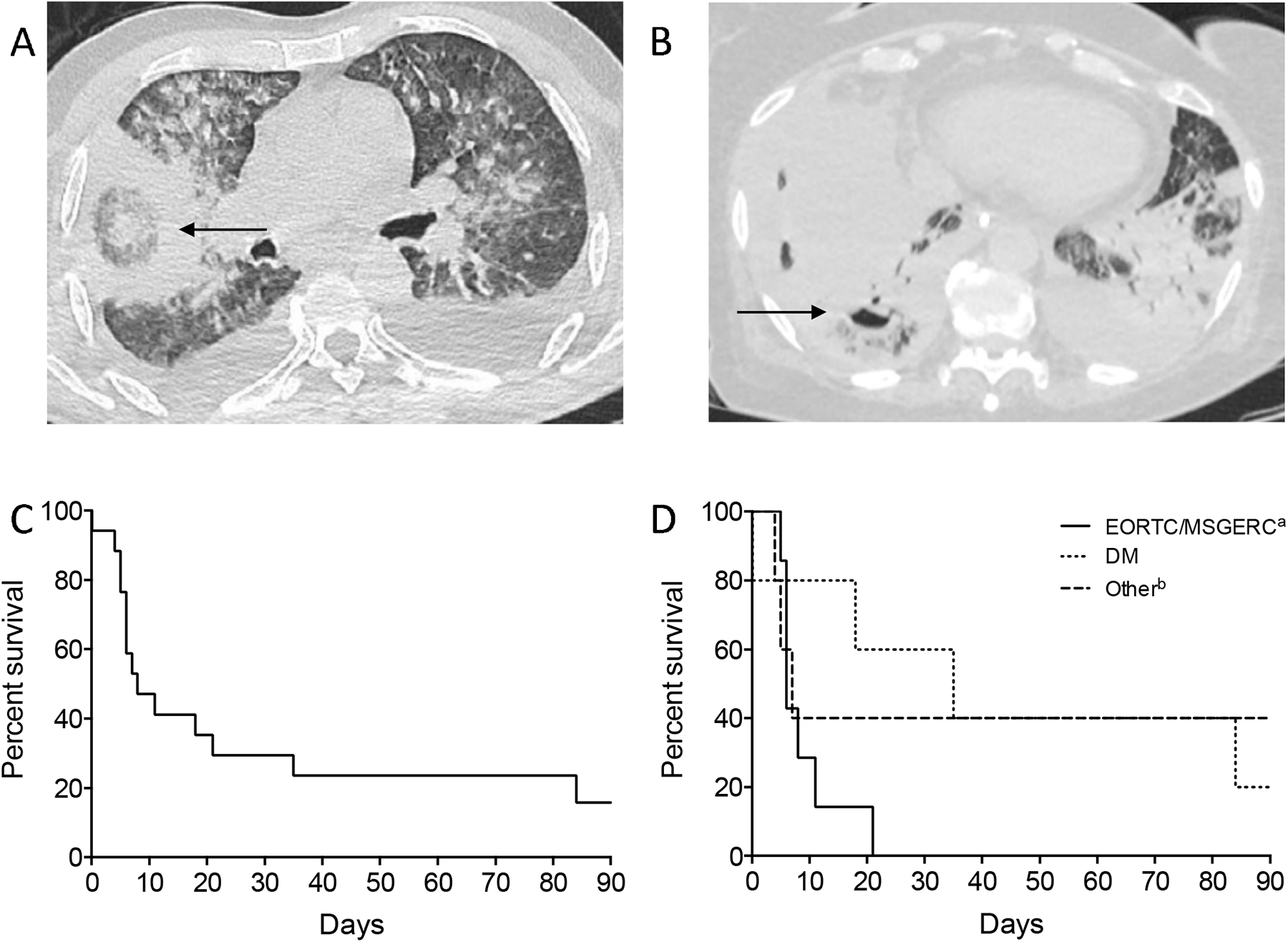
COVID-19 associated mucormycosis in France. A: Chest CT scan showing a reversed halo sign. B: Chest CT scan showing a cavitation in a consolidation. C: Twelve-week survival after COVID-19 associated mucormycosis diagnosis* (n=17). D: Twelve-week survival according to the host factors. * Day 0 corresponds to the first positive sample for mucormycosis. One patient was alive after 43 days but had not yet twelve weeks of follow-up. ^a^Classical EORTC/MSGERC criteria were considered before COVID-19. ^b^Other: five patients receiving corticosteroids for COVID-19 (n=3) or without risk factors (n=2). DM: diabetes mellitus.

### Mycology

CA-Mucor was classified as proven in five (29%) patients and probable in twelve (71%). The culture grew *Mucorales* in samples from eleven (65%) patients (bronchoalveolar lavage (BAL, n=5), tracheal aspirate and biopsy (n=3, each)). *Mucorales* PCR assay (adapted from [7] in seven centers, from [8] in one center, and from MycoGENIE®, Ademtech, France, in one center) was found positive in fifteen (88%) patients [serum (n=14), BAL (n=7), tissues (n=3), peritoneal fluid (n=1)]. Ten patients had more than one positive sample (on different site). PCR was the only means of diagnosis for four patients including two with positive serum and BAL samples. Histology identified hyphae compatible with *Mucorales* in biopsies from four patients.

*Mucorales* was identified to the genera or species level by culture or species-specific PCR in fourteen (82%) patients mainly (n=9, 64%) with *Rhizopus* [*Rhizopus microsporus* (n=6; 43%), *Rhizopus delamar* (n=2; 14 %) and *Rhizopus arrhizus* (n=1; 7%)], secondary (n=4, 29%) with *Rhizomucor* [*Rhizomucor pusillus, Rhizomucor miehei* (1 each, 7%)], and with *Lichtheimia* sp. in only one case (7%). All species identified from both culture and PCR were concordant. The cases of *Rhizomucor* occurred in three patients with HM and in one patient with pulmonary carcinoma.

### Other fungal infections

Five (29%) patients developed COVID-19 associated aspergillosis (CAPA), a median of two days [-28–0] before CA-Mucor. All patients with CAPA and CA-Mucor died before week 12 of mucormycosis. One (6%) patient had invasive candidiasis concomitantly to CA-Mucor.

### Treatment and outcome

Five (29%) patients died before receiving any specific treatment. Twelve (71%) patients were prescribed liposomal amphotericin B (n=10, 59%), or isavuconazole (n=2, 12%). Three (18%) patients had surgery, two for rhino-orbito-cerebral mucormycosis and one for colonic perforation.

Global mortality was 76% (13/17) at week 6 and 88% (14/16) at week 12 (figure 1C and D). One patient was alive after 43 days but had not yet 12 weeks of follow-up. Death occurred after a median of 34 days [15-119] after the first symptoms of COVID-19 and after a median of 7 days [0-84] after the first positive sample for CA-Mucor.

## DISCUSSION

We here reported seventeen cases of CA-Mucor in France, the largest series from one country outside India [4,9]. We observed a large spectrum of clinical presentations and of host factors, and evidenced a high mortality of >75% by 6 weeks. These findings differ from our historical (2005-2007) series of 101 mucormycoses in France (RetroZygo, [10]) and from CA-Mucor reported in India [4,9,10].

If we compare mucormycoses risk factors in France from present CA-Mucor series and RetroZygo series, classical EORTC/MSGERC host factors were recorded in 41% *vs*. 52%, respectively, and DM in 47% *vs*. 32% [5,10]. The frequency of underlying DM was far lower than that recorded in CA-Mucor in India (60% to 95%) [4,9]. By contrast, the frequency of hematological malignancy in CA-Mucor was higher compared to India (35% vs 1%) [9].

Clinical spectrum was also different, with more frequent pulmonary (53% *vs*. 28%), and less frequent rhino-orbito-cerebral locations (12% *vs*. 25%) in the current series *vs*. historical RetroZygo Study. The presentation differed clearly from India, where CA-Mucor mainly presents with rhino-orbito-cerebral locations (>80%) [9]. This difference could be explained by higher prevalence of DM in Indian patients.

Culture and/or histology were positive in 76% of patients, while diagnosis of CA-Mucor was based only on a positive Mucorales PCR in four patients (24%). The broad use of *Mucorales* PCR could partly explain the higher frequency of CA-Mucor in France compared to other European countries [3,7]. *Rhizopus microsporus* was the most frequent species in this small series. It is likely though that *Mucorales* being present in the environment, the species recovered are influenced by the geographic area, as well as the anatomic site and the underlying risk factors explaining the differences in species distribution among studies independently of the COVID context [4].

Twelve week mortality was very high (88%) in our current CA-Mucor study, by comparison with RetroZygo Study (44%), and with CA-Mucor in India (40-50%) [4,9,10]. It is also higher than what is reported for CAPA [1,11]. This major difference might be partly explained by the higher frequency of pulmonary or disseminated presentations, which are classically associated with a poorer prognosis compared with rhino-orbito-cerebral locations. The severity of COVID-19 itself and the high proportion of patients hospitalized in the ICU might also account for these differences.

The differences in comorbidities, anatomical location and prognosis between our current series and CA-Mucor in India might be explained by a higher frequency of patients with DM in India and the higher burden of mucormycosis, as well as by differences in the management of patients in ICU between the two countries [12].

Despite the multicenter design, the limitations of our study are the small number of cases and the retrospective design. However, epidemiological surveillance in France is based on a reliable and sustained collaboration between French mycologists and the NRCMA. This makes unlikely a major erroneous reporting.

CA-Mucor has a high mortality in this study. Better knowledge, identification and earlier treatments of CA-Mucor might help to improve the prognosis. International studies are warranted to better understand and assess CA-Mucor.

## Data Availability

The data that support the findings of this study are available on request from the corresponding author

